# Modular coupling of structure-function reveals network integration (rather than segregation) as the key mechanism for cognitive task discrimination

**DOI:** 10.1101/2025.03.10.25323674

**Authors:** Izaro Fernandez-Iriondo, Antonio Jimenez-Marin, Naiara Aginako, Gorka Zamora-López, Asier Erramuzpe, Paolo Bonifazi, Jesus M. Cortes

**Affiliations:** Computer Science and Artificial Intelligence, University of the Basque Country (UPV/EHU), San Sebastian, Spain; Computational Neuroimaging Lab, Biobizkaia Health Research Institute, Barakaldo, Spain; Informatics Engineering Doctoral Programme, University of the Basque Country (UPV/EHU), San Sebastian, Spain; Center for Brain and Cognition, Universitat Pompeu Fabra, Barcelona, Spain; IKERBASQUE: The Basque Foundation for Science, Bilbao, Spain; Department of Cell Biology and Histology, University of the Basque Country (UPV/EHU), Leioa, Spain; Department of Physics and Astronomy, University of Bologna, Bologna, Italy

## Abstract

Understanding how structural and functional brain networks interact to support cognitive processes remains a central challenge in systems neuroscience. In this study, we investigate the dynamics of structure-function coupling (SFC) at the modular level across different cognitive tasks using multimodal neuroimaging data, including anatomical, diffusion, functional at rest and functional at different tasks. By constructing high-resolution structural and functional connectivity matrices, we assessed intra-modular (SFC-INT) and inter-modular (SFC-EXT) coupling to examine their roles in task-specific reorganization. Our results reveal that variations in SFC during cognitive tasks are primarily driven by changes in inter-modular coupling, emphasizing network integration over segregation. Specifically, tasks demanding higher cognitive flexibility, such as the gender stroop task, exhibited increased SFC-EXT, indicating enhanced integration between modules. In contrast, tasks focused on memory processing showed a tendency toward segregation, with lower SFC-EXT values. These findings highlight the significance of inter-modular integration as a flexible and dynamic mechanism underlying cognitive task discrimination. Our study advances the understanding of modular brain network dynamics, suggesting that the brain’s ability to integrate information across modules plays a pivotal role in cognitive flexibility and task performance.

## 1 Introduction

Understanding the relationship between different types of brain connectivity is a key challenge in systems neuroscience [1]. Structural connectivity (SC), defined by white matter pathways, provides the foundation for neuronal communication, while functional connectivity (FC), measured by the statistical similarity of neural activity, reflects the brain’s dynamic interactions [2–4]. A central question in this field is how these two connectivity types interact to support cognitive processes and adapt to task demands [5–8].

It is well established that SC constrains FC at rest, forming large-scale networks that remain stable across different states [9]. However, increasing evidence suggests that SC-FC relationships change dynamically during cognitive tasks, reflecting the brain’s ability to reorganize [10–12]. The extent and nature of this reorganization remain open questions, particularly regarding the balance between network segregation (local specialization) and integration (distributed processing) [13, 14].

In this study, we analyze how modular structure-function coupling (SFC) varies across cognitive tasks, focusing on two key levels. At the global level, we assess how task execution impacts overall SFC, revealing that inter-modular coupling (SFC-EXT), which measures the integration of modules with the rest of the network, exhibits the greatest variation across tasks. This suggests that cognitive flexibility is primarily driven by changes in integration rather than segregation. At the modular level, we explore how SFC reorganization aligns with task-specific demands, observing that each cognitive task induces distinct patterns of modular reorganization, with some favoring integration while others show increased segregation.

These findings emphasize the importance of modular reorganization in brain network dynamics and support the idea that integration, rather than segregation, plays a central role in discriminating cognitive tasks. By leveraging a multimodal approach and high-resolution parcellation, our study advances the understanding of how structural and functional networks interact flexibly to meet changing cognitive demands. Future research should further explore how these mechanisms contribute to cognitive flexibility and task performance across different domains.

## 2 Materials and Methods

### 2.1 Participants

A total of 216 participants from the Amsterdam Open MRI Collection (AOMIC) dataset [15] were initially considered in this study accompanied by demographic and psychometric information. This data is freely accessible via OpenNeuro data-sharing platform and specifically here, we made use of the PIOP1 dataset which includes T1w, DWI, resting-state fMRI and fMRI images while performing several cognitive tasks per each participant [16]. For this study, we selected the following tasks: emotion matching, working memory, emotion anticipation, and gender stroop. After data quality assurance (see the Supplementary Material), the final number of included participants was *P*= 54 with ages ranging from 18.25 to 26.0 years (mean 22.22, standard deviation 1.61).

### 2.2 Cognitive tasks

In this study, four task-related functional data sessions were performed per participant. The **emotion anticipation task** examined the ability to anticipate and respond to emotions, focusing on curiosity and prediction; participants were initially given cues predicting the likelihood of viewing an emotionally negative or neutral image, followed by the actual presentation of the image, testing anticipatory emotional responses through 30 unique images. The **emotion matching task**, inspired by earlier paradigms from Hariri et al. [17], assessed facial emotion processing by requiring participants to match the emotional expression of a target face to one of two probe faces. The **gender stroop task** measured cognitive conflict and control using a face-gender variant of the stroop test, where participants were presented with congruent or incongruent face-label pairs and asked to identify the gender of the face while disregarding the distractor word. Lastly, the **working memory task** aimed to assess how participants maintained and manipulated visual information in both active and passive conditions, responding to orientation changes or maintaining attention according to specific cues, encompassing various stages of working memory processing. For a detailed description of the methodologies see the full article [15].

### 2.3 Imaging data acquisition

#### Anatomical data

A high-resolution T1-weighted image was acquired with a 3D magnetization prepared rapid acquisition gradient echo (3D MPRAGE) with parameters: TR/TE (ms) = 8.5*/*3.9, voxel size = 1 × 1 × 1 mm^3^, field of view (FOV) = 188 × 240 × 220 (RL × AP × FH ^1^; mm), flip angle = 8*^◦^*and a duration of 6 min 3 sec.

#### Diffusion weighted imaging (DWI)

A spin-echo diffusion weighted imaging (SE-DWI) sequence was acquired with the following parameters: TR/TE = 7456/86, voxel size = 2 × 2 × 2 mm^3^, slice thickness = mm, field of view (FOV) = 224 × 224 × 120 (RL × AP × FH; mm), number of *b*_0_ images = 1, number of diffusion-weighted directions = 32, DWI b-value = 1000 s/mm^2^, matrix size = 112 × 112, number of slices = 60, slice gap = 0 mm, flip angle = 90*^◦^* and a duration of 5 min 27 sec.

#### Resting functional data

Acquired with a gradient echo EPI scan technique using the following parameters: TR = 750 ms, TE = 28ms, FOV (RL × AP × FH; mm) = 240 × 240 × 118 mm^3^, voxel size = 3 × 3 × 3 mm^3^, matrix size = 80 × 80, number of slices = 36, slice gap = 0.3mm and flip angle = 60*^◦^*.

#### Task functional data

Acquired with a gradient echo EPI scan technique using the following parameters: TR/TE = 2000, FOV (RL × AP × FH; mm) = 240 ×240 ×122 mm^3^, voxel size = 3 ×3 ×3 mm^3^, matrix size = 80 ×80, number of slices = 37, slice gap = 0.3mm and flip angle = 76.1*^◦^*.

### 2.4 Imaging data preprocessing

#### Diffusion images

Preprocessed using a custom pipeline making use of brain-extracted T1 images [18, 19], MRtrix3 (v3.0 RC3) [20], FSL (v6.0.1) [21], and ANTs (v2.3.1). Our DWI pipeline involved cleaning the raw images using *DWIdenoise* [20, 22–24] and *DWIpreproc* [20, 25–27] tools. Next, we performed whole-brain probabilistic tractography using the iFOD2 [28] tracking algorithm in MRtrix3 [20]. The tractography was initiated from seeds located at the gray matter-white matter interface, with a selection of 3 million fibers based on an angle threshold of *<*45*^◦^*and a length threshold of *<*200 mm, following the recommendations in the MRtrix3 documentation. The tool *SIFT2* [29] was used to correct for the effect of crossing and disconnection of fibers on the quantification of the total number of streamlines.

#### Functional images

Preprocessed with our pipeline previously used in [18,19] including slice-time correction, and alignment of volumes for head-motion artifact correction with MCFLIRT (FSL 6.0.1) [30–32]. Movement time-courses obtained from MCFLIRT were used as an input of ICA-AROMA tool [33], a data-driven method used to identify motion-related independent components (IC) in fMRI data. After intensity normalization, time-courses of the motion-related ICs, the 5 PCA components of the cerebrospinal fluid and the 5 components of the white matter signals with more variance explained, and linear and quadratic trends were removed together with a 0.01–0.08 Hz band-pass. In the case of the task fMRI, the design event matrix was also removed. Finally, images were normalized to the MNI152 brain template (3 × 3 × 3 mm^3^ voxel size) and spatially smoothed with a 6 mm full width at half maximum isotropic Gaussian kernel.

### 2.5 Parcellation of neuroimaging data

In our approach, we first delineated a functional partition comprising a large number of micro-regions (*>* 2000), that lately will represent the different nodes in SC and FC. We employed an unsupervised clustering method that groups voxels into the same region based on voxel-level dynamics similarity [34]. In an effort to integrate anatomical constraints among these micro-regions, our previous work involved various, independent, functional partitions across various macro-regions [35]. Here, we considered the following eight macro-regions: *frontal lobe, parietal lobe, occipital pole, temporal pole, insula, brain stem, cerebellum, and subcortical structures* (the latter achieved by pooling together the thalamus, caudate, putamen, pallidum, amygdala and hippocampus). Therefore, although we used a functional partition, our approach ensures that the derived functional partition is informed by, and reflective of, the brain’s intrinsic anatomical macro-organization.

### 2.6 Population average connectomes

In our methodology, we obtained subject-specific SC and FC matrices. Subject-level SC matrices were obtained by counting the number of white matter streamlines connecting all possible pairs of micro-regions. For FC, we computed a time series for each of the micro-regions, by averaging within micro-region voxel time-series, and then assessed the statistical dependence between all pairs of micro-regions using the Pearson correlation coefficient. The next step is to build population pSC and pFC matrices, that here was done by calculating for each link in pSC and pFC, the median value of all links across all individuals. Finally, out of the original number of micro-regions, we removed those exhibiting null connectivity values to any other region in the pSC. For details on code to generate these steps, see https://github.com/compneurobilbao/bha2.

### 2.7 Evaluating modular organization

We identified modules or communities in both pSC and pFC, with the purpose of using as a reference the modules in one modality for comparison with the other (ie., comparing structural modules to functional ones and vice versa). While it is true that the most celebrated method for community detection is modularity maximization [36], it has been acknowledged by previous authors that modularity maximization encounters a variety of serious conceptual flaws [37, 38]. The most problematic one is that it always finds high-modularity partitions in networks sampled from its own null model [37], providing a hard interpretation to the communities found at the maximum of modularity. A different strategy to modularity maximization that sorts out this problem is Bayesian Stochastic Block-Modeling [39], but this alternative needs for prior information before Bayesian Inference, which is also unknown for many practical situations.

Here, and following previous work [35, 40], we decided to perform hierarchical agglomerative clustering, and then calculate Newman’s modularity *Q* at all levels of the tree [41]. The final list of modules is chosen at the dendrogram level with highest *Q*, for both pSC and pFC. Therefore, we do not search for the optimal partition that maximizes *Q*; instead, among different hierarchical partitions, we use the representation with highest *Q*. This was done for pSC and pFC. Similar to [35], before hierarchical agglomerative clustering in pFC, we thresholded the values of the matrix to achieve the same density of non-zero links as in pSC. Moreover, we calculate the correlation distance between pairs of nodes using the (1 *−* cosine(pFC)) matrices from the functional data and pSC from the structural data, followed by hierarchical agglomerative clustering based on the previous distances between regions.

### 2.8 Assessment of modular SFC

The functional implications of every structural module is evaluated through the SFC which was assessed by the Pearson correlation coefficient *r* as a similarity measure:

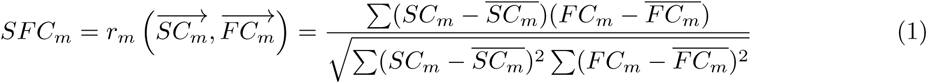

where 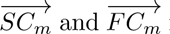 represent the vector-wise representations of connectivity matrices SC*_m_*and FC*_m_* corresponding to the module *m*.

A module *m* is composed of a set of *n* nodes, and its structural connectivity matrix SC*_m_* and functional connectivity matrix FC*_m_* are of size *n×N*, where *N* represents the total number of regions of interest (ROIs) in the brain. The connectivity within the module (internal connectivity) is represented by the *n × n* submatrix corresponding to the block on the diagonal of SC*_m_* and FC*_m_*. Conversely, the external connectivity of module *m* refers to the submatrix of size (*N − n*) *× n*, which captures the interactions of module *m* with the rest of the brain. This decomposition into internal and external connectivity is illustrated in Fig. 1, which provides a clearer representation of our methodological approach.

**Figure 1:**
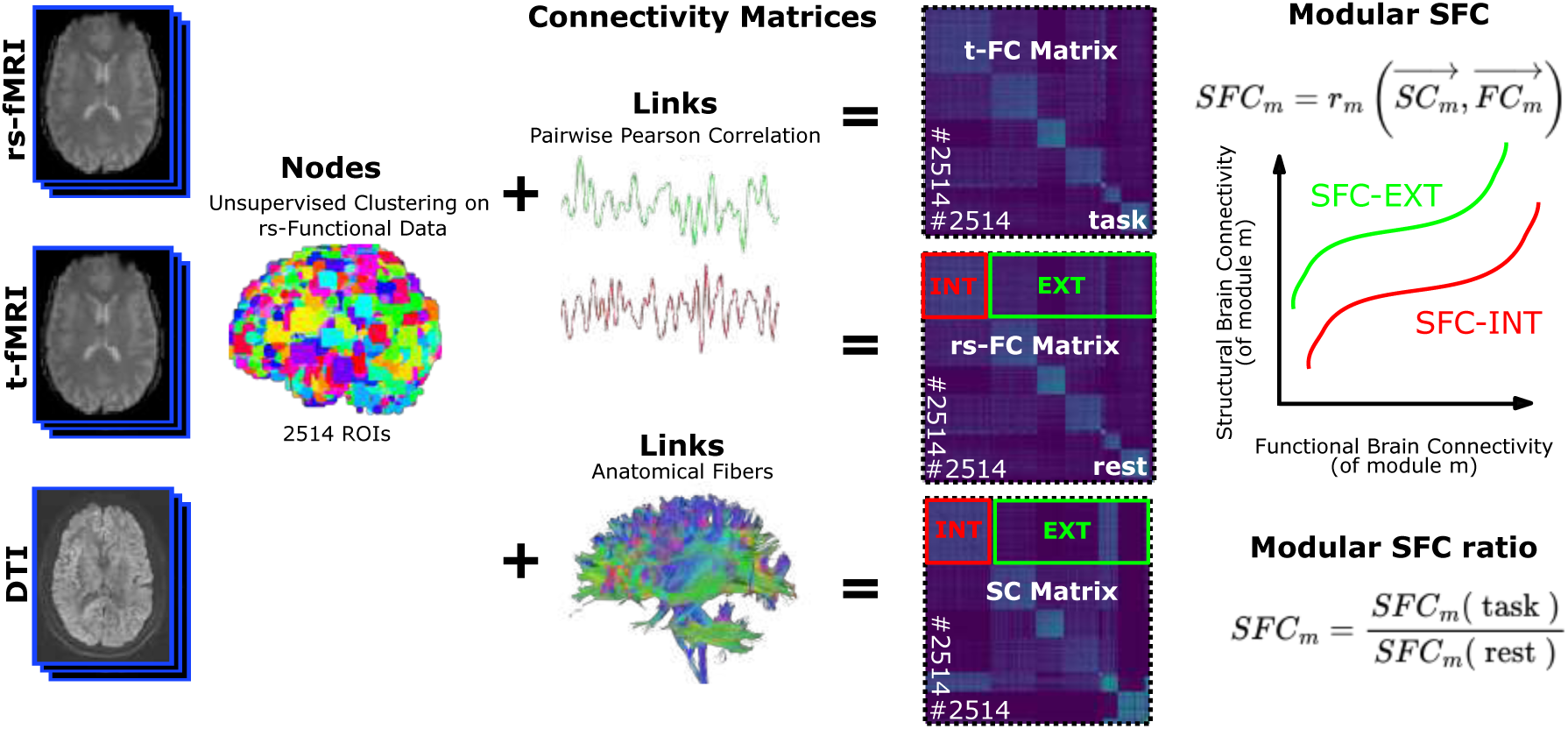
Methodological pipeline. This figure illustrates the multimodal neuroimaging approach used to analyze structural-functional coupling (SFC) in the human brain. Three imaging modalities are used: resting-state functional MRI (rs-fMRI), task-based functional MRI (t-fMRI), and Diffusion Tensor Imaging (DTI). Nodes are defined through unsupervised clustering of rs-fMRI data, identifying 2514 regions of interest (ROIs). Functional connectivity (FC) is computed using pairwise Pearson correlations of BOLD signals, while structural connectivity (SC) is based on DTI-derived anatomical fibers. These connectivity matrices—SC, resting-state FC (rs-FC), and task-based FC (t-FC)—serve as the basis for SFC computation. To assess modular SFC, hierarchical agglomerative clustering is applied separately to the populational resting-state FC matrix and the SC matrix, allowing the identification of structural and functional modules. SFC is then computed separately for intra-modular (SFC-INT) and inter-modular (SFC-EXT) connectivity. Finally, the modular SFC ratio is derived by comparing SFC during tasks to SFC at rest, providing insight into task-related reorganization of brain connectivity.

**Figure 2:**
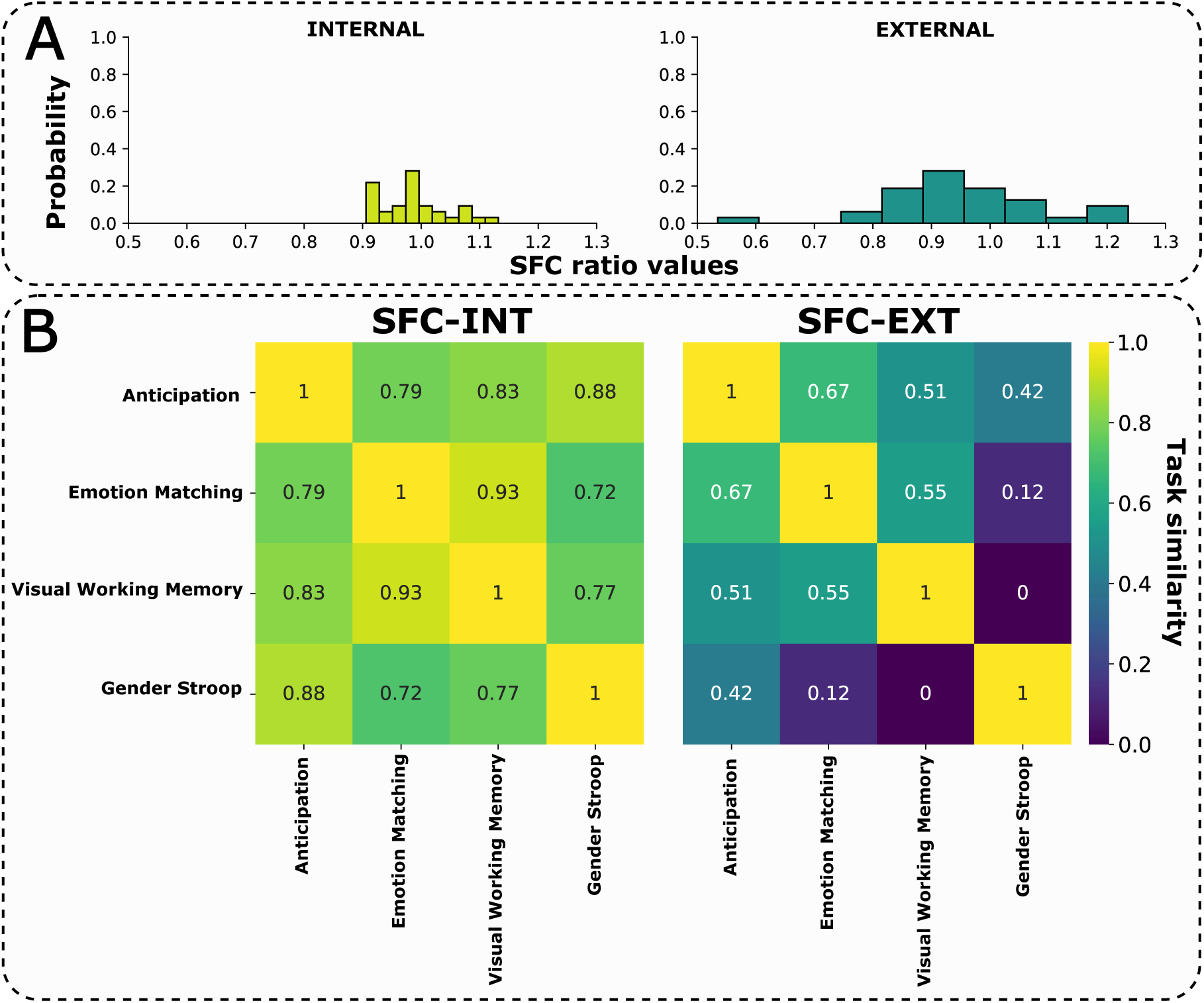
Distribution of SFC ratios and task similarity for the structurally driven partition. **A) Distribution of SFC Ratios for Internal and External Connectivity.** The histograms represent the probability distribution of SFC ratio values for internal (left) and external (right) connectivity. These SFC ratios serve as the basis for computing task distances, which are analyzed in the following panel. **B) SFC Similarity Between Tasks.**Each element in the heatmaps represents the SFC similarity (1 - SFC distance) between pairs of tasks. The left heatmap shows intra-modular SFC (SFC-INT) similarity, a metric for task segregation, while the right heatmap displays inter-modular SFC (SFC-EXT) similarity, used here as a metric for network integration. These similarities were calculated for the structural partition, and similar results were found for the functional partition (see Figure S1).

The modular SFC of a given module *m* can be assessed both within the module itself (SFC-INT) and in relation to its connections with the rest of the modules (SFC-EXT). The **SFC-INT** is defined as 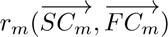, where *m* = 1*,…, M*, which captures the similarity within each module (internal connectivity). Conversely, the **SFC-EXT** accounts for the similarity in the links connecting one module to the rest of modules, expressed as 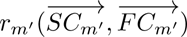, where *m* represents the external connectivity [42–45] of module *m*.

To assess the interplay between SFC-INT and SFC-EXT, we analyzed their relative changes across tasks; to do so, both metrics were normalized by their resting-state values to capture task-induced deviations. Extreme cases emerge based on the relative values of SFC-INT and SFC-EXT. When both exceed 1, the task is structurally driven, indicating that internal and external functional connections adhere more closely to the brain’s structural architecture. Conversely, when both are below 1, the task is functionally driven, reflecting greater flexibility as connections deviate from structural constraints. Modules exhibiting intermediate values represent a balance between these two extremes, maintaining a dynamic equilibrium between structural fidelity and functional adaptability.

This framework allows for the characterization of brain module dynamics along a continuum, where tasks promote segregation, reinforcing stable internal connectivity; integration, enhancing flexible intermodular communication; or an intermediate state, balancing both processes.

### 2.9 Task distance computation at modular level

To analyze how the SFC metric varies from one task to another at a global level—without delving into modular details—we compute the distance between tasks. This allows us to quantify their dissimilarity based on the modular SFC representation. The distance is calculated separately for both internal coupling (SFC-INT) and external coupling (SFC-EXT) to capture distinct aspects of modular interactions.

The distance between two tasks, in terms of the SFC metric, is defined as the Euclidean distance between their respective modules:

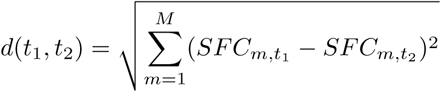

Where:

- *t*_1_ and *t*_2_ represent two distinct tasks.
- *SFC_m,t_*_1_ and *SFC_m,t_*_2_ are the values of the SFC metric for module *m* in tasks *t*_1_ and *t*_2_, respectively.
- *m* spans from 1 to *M*, the total number of modules resulting from community detection.
- *d*(*t*_1_*, t*_2_) is the Euclidean distance between tasks *t*_1_ and *t*_2_ in the space defined by the SFC metric.

To construct the distance matrix between tasks, *d*(*t*_1_*, t*_2_) is computed for each pair of tasks, enabling the visualization of their differences through a heatmap. By computing this distance separately for SFC-INT and SFC-EXT, we can distinguish how task-driven changes impact internal modular cohesion versus inter-modular interactions.

## 3 Results

A population of *P* = 54 healthy participants with quality-assured neuroimaging data was studied here, including anatomical imaging, diffusion imaging, resting-state functional imaging, and task-based functional imaging during four different cognitive tasks (emotion anticipation, emotion matching, gender stroop, and working memory). Prior to assessing the modular structure-function coupling (SFC), we obtained population matrices pSC and pFC^rest^ with dimensions 2482×2482. Before applying hierarchical agglomerative clustering (HAC), pFC^rest^ was thresholded by progressively removing the lowest values in —pFC^rest^— until the density of non-zero links matched that of pSC, which in our data was equal to 8.34%.

Next, we applied HAC to both pSC and pFC^rest^, and evaluated the modularity Q across different levels of the resulting dendrogram. The maximum Q values obtained yielded M = 8 non-overlapping structural modules with a modularity index Q = 0.74, and M = 14 functional modules with Q =

0.47. The final sets of structural modules and functional modules are depicted in Fig.3 and Fig.4, respectively. The major methodological steps and the various measures used are summarized in Fig.1. Our primary objective was to examine how structural connectivity reorganizes during different cognitive tasks and to assess the dynamic relationship between SC and FC under varying conditions. To this end, we computed two key metrics: intra-modular structure-function coupling (SFC-INT), which quantifies the alignment between SC and FC within each module, and inter-modular structure-function coupling (SFC-EXT), which measures the coupling between a module and the rest of the brain (see Fig. 1).

**Figure 3:**
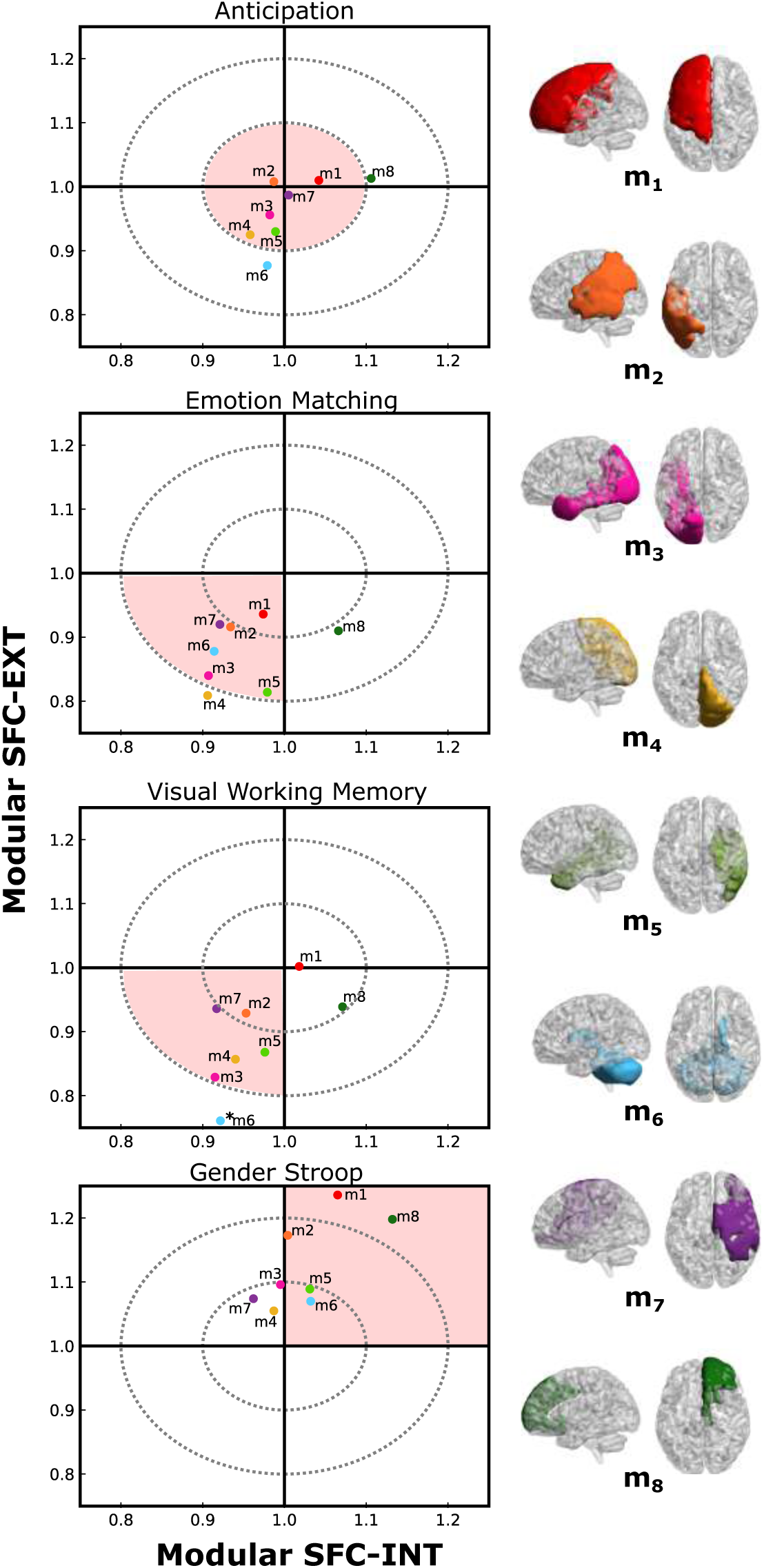
Visual representation of different tasks for structurally-defined modules. The x-axis represents intra-modular SFC, while the y-axis displays the inter-modular SFC ratio (with respect to resting). Each row corresponds to a different cognitive task, and each point in the bidimensional space represents a structural module shown on the right panel.

**Figure 4:**
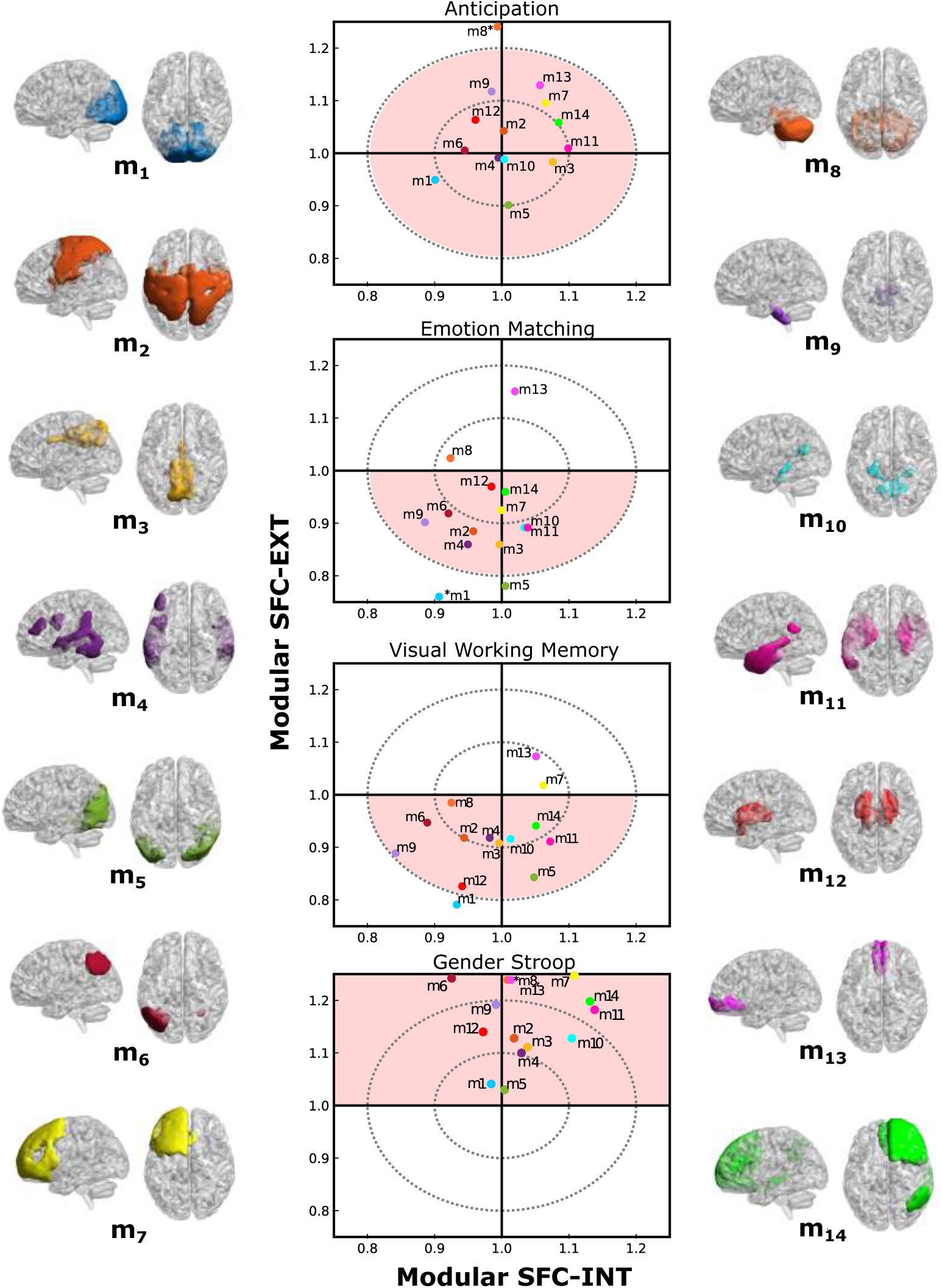
Visual representation of different tasks for functionally-defined modules. Similar to figure 3, we represent inter-modular versus intra-modular SFC ratios for the different functional modules and across different tasks.

To better understand the differences in task-dependent SFC, we plotted the probability distribution of SFC ratio values separately for internal and external connectivity (see Fig. 2A). The distribution of SFC-INT values appears narrower compared to that of SFC-EXT, suggesting a more constrained variability in the internal connectivity domain. In contrast, the broader distribution observed in SFC-EXT indicates greater variability across tasks, which aligns with principles of signal detection theory: a wider distribution allows for higher discriminability between conditions. This suggests that distinguishing tasks based on SFC-INT may be more challenging due to its restricted range, whereas the broader spread of SFC-EXT values enhances its potential for task differentiation.

These differences in variability are further reflected in the task similarity analysis (see Fig. 2B). Task differences are more pronounced for SFC-EXT, which reflects integration, than for SFC-INT, which reflects segregation. This indicates that while the intrinsic alignment between structural and functional connectivity within modules remains relatively stable, the capacity of these modules to integrate with the broader network is highly sensitive to cognitive demands. In essence, integration processes appear to be more flexible and task-dependent than segregation processes. The same pattern is observed for the partitioning of functional modules (see Fig. S1), where the stability of segregation contrasts with the dynamic nature of integration under different task conditions.

At the modular level for the structurally driven partition (see Fig. 3), the patterns of SFC reveal distinct shifts across tasks. In the emotion anticipation task, modules exhibit a balanced coupling between SFC-INT and SFC-EXT, clustering near the center of the plot. However, during emotion matching and visual working memory tasks, most modules display a segregation-dominant pattern, with both SFC-INT and SFC-EXT values below 1. Conversely, the gender stroop task shows a marked shift toward integration, as most modules occupy the quadrant where both SFC-INT and SFC-EXT values exceed 1.

A similar pattern emerges for the functional modules (see Fig. 4). Regardless of the partitioning method, functional modules consistently show balanced coupling in the emotion anticipation task, a shift toward lower SFC-INT and SFC-EXT values in the emotion matching and visual working memory tasks, and a marked increase in both metrics during the gender stroop task. This consistency across both structural and functional partitions underscores that each cognitive task imposes a distinct balance between segregation and integration, with SFC-EXT being particularly sensitive to the task demands.

## 4 Discussion

In this study, we explored the extent to which structural connectivity supports brain function by assessing structure-function coupling (SFC) at the modular level and its variation across different cognitive tasks. By constructing high-resolution networks encompassing the entire brain, including the cortex, subcortical structures, and cerebellum, we identified data-driven structural and functional modules to evaluate modular SFC. This methodological framework allowed us to systematically map functional and structural connectivity through SFC across different cognitive tasks, revealing distinct patterns of modular behavior. Notably, our results demonstrate that analyzing SFC at the modular level preserves functional specificity, allowing for a more precise differentiation between tasks. This approach enhances the detection of task-specific connectivity patterns by focusing on intra- and inter-modular coupling, which provides a more accurate representation of neural organization. By maintaining the integrity of structural and functional modules, modular SFC analysis captures meaningful variations in connectivity, offering a powerful strategy for understanding brain function in different cognitive contexts.

Our findings indicate that task-dependent variations in SFC are primarily driven by changes in inter-modular coupling (SFC-EXT), which reflects network integration, rather than intra-modular coupling (SFC-INT), which represents segregation. This suggests that while the intrinsic alignment between SC and FC within modules remains relatively stable, cognitive demands modulate the brain’s ability to integrate across different modules. Specifically, the emotion anticipation task exhibited a balanced coupling profile across modules, while tasks such as emotion matching and visual working memory showed a shift toward increased segregation. In contrast, the gender stroop task was characterized by enhanced integration, emphasizing the task’s reliance on flexible, distributed processing.

These findings align with previous research demonstrating task-dependent variations in functional organization [10–12, 12, 46–51]. Notably, our results (see fig. S2) corroborate prior evidence that high-attention tasks, such as gender stroop, typically involve reductions in functional activity, while memory-intensive tasks, such as visual working memory and emotion matching, show substantial increases in functional connectivity [46] relative to rest. The emotion anticipation task appears to fall between these cognitive domains, as it involves both memory and attention processes. This pattern reflects a trade-off between segregation and integration, whereby attentional tasks promote more efficient, localized processing, whereas memory tasks require broader, distributed interactions.

Furthermore, our results (see fig. S3) support the notion that primary sensory and motor areas exhibit stronger SFC coupling than transmodal areas, which are more involved in integrative and executive functions. Consistent with Bassett’s findings, we observed higher coupling indices in unimodal regions such as the occipital and parietal areas, whereas subcortical and frontal regions exhibited lower coupling, reflecting their distinct roles in cognitive processing [52–57].

By leveraging a multiscale and multimodal approach, we demonstrated that analyzing brain networks at a modular level—rather than at a lower regional scale—provides a more comprehensive understanding of network interactions. Our mapping of different tasks into the space of intra- and inter-modular interactions revealed that inter-modular SFC, in particular, serves as a key marker for differentiating task-specific reorganization. This approach captures the dynamic and heterogeneous nature of network interactions during different cognitive tasks, highlighting modular reorganization rather than specific module-task associations.

In summary, our results show that brain network integration is more flexible and adaptable than segregation, playing a key role in responding to different cognitive tasks. Future studies should focus on how these inter-modular connections adapt to various cognitive demands to gain a deeper understanding of brain function

## Supplementary Material

### Data quality assurance

The data were preprocessed for N=216 participants, and after, we applied a exclusion criteria based on well-established image quality standards (motion quantity threshold equals to an average FD*<*0.5), complete cerebellar acquisition, and well-aligned co-registrations between T1w and functional images. Moreover, to ensure accurate data preprocessing, we employed a graphical strategy analogous to that of the Brain Connectivity Toolbox [58].

Specifically, following standard preprocessing steps such as brain denoising, ensuring stationarity, filtering, and regressors removal (for details see Image preprocessing section), the probability distributions of the functional connectivity values between voxel pairs should be centered around zero, meaning the mean of the FC distribution should ideally be zero post-processing. However, in our study, each participant underwent resting-state scans plus four different tasks. To guarantee robust preprocessing across all five conditions simultaneously, we initially constructed five-dimensional vectors (each component corresponding to the mean FC value for each condition), and then ensured that their norm was less than 0.35. This approach enabled us to rigorously select *P* = 54 participants based on stringent imaging criteria. Ultimately, our analysis is based on population-level matrices derived from median values among these 54 participants, thereby providing a sample size more than sufficient for this type of methodological strategy. Participant’s age ranged between 18.25 and 26.0 years (mean 22.22 and standard deviation 1.61).

## Data Availability

All data produced are available online via OpenNeuro data sharing platform

https://openneuro.org/datasets/ds002785/versions/2.0.0

---

**Figure S1:**
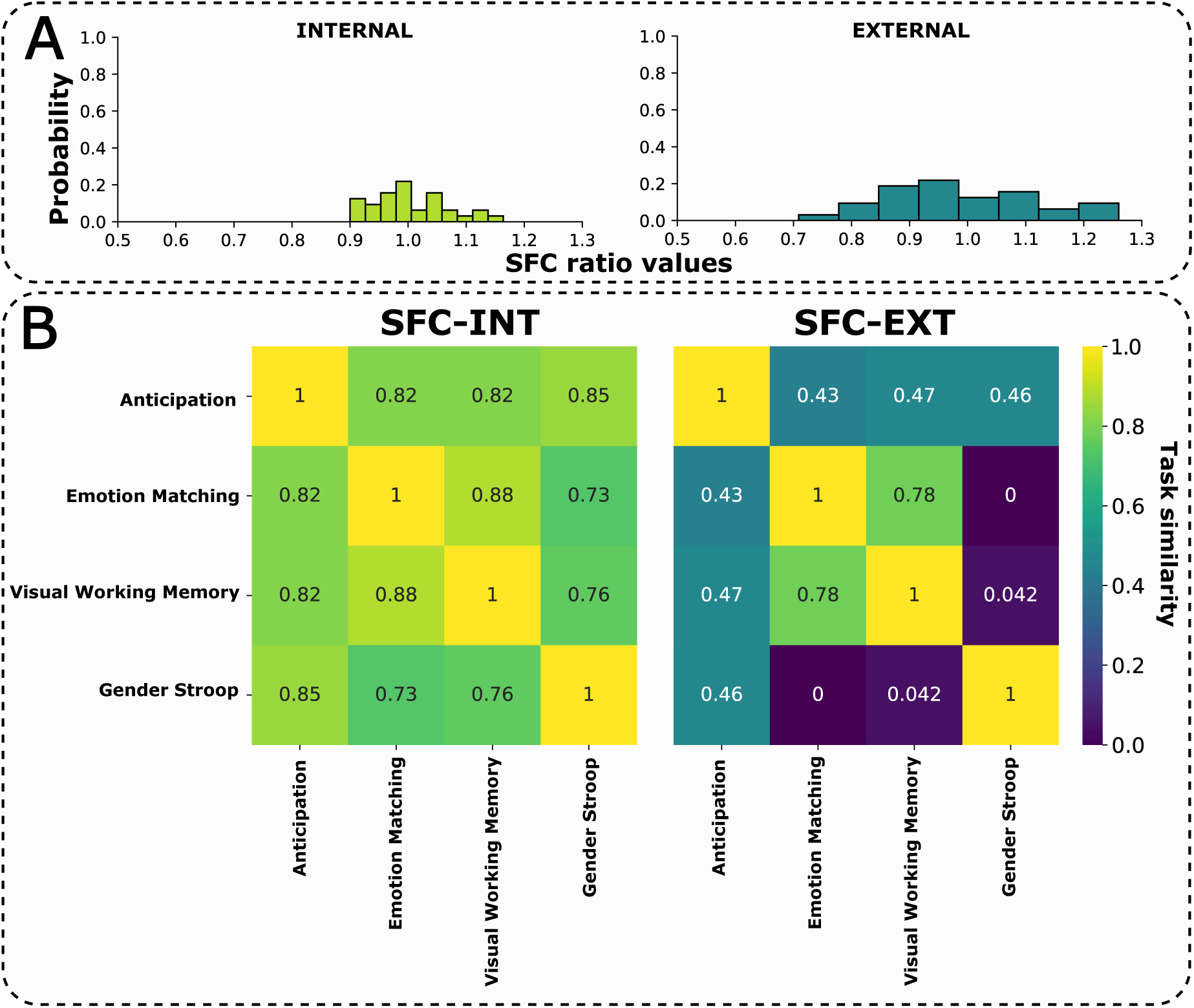
Distribution of SFC ratios and task similarity for the functionally driven partition. Similar to figure 2 SFC ratios distribution on panel A and SFC simialrity between tasks on panel B.

**Figure S2:**
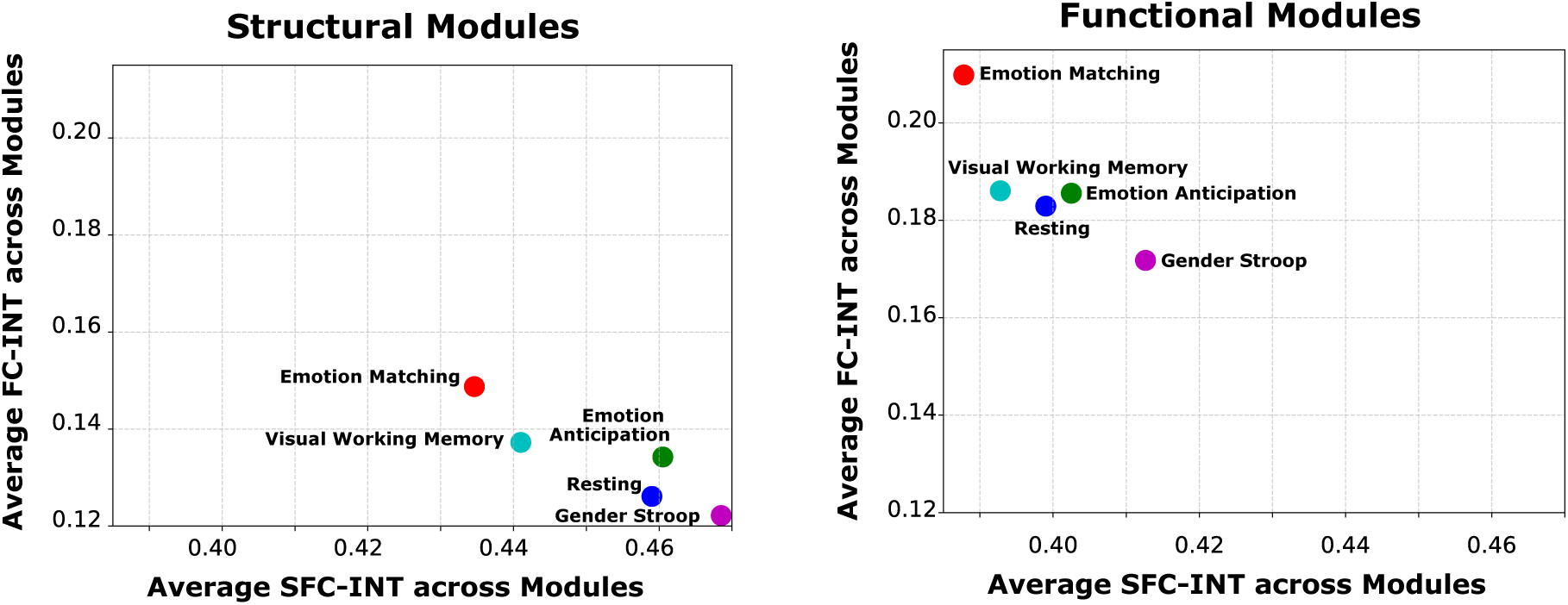
Scatter plots showing the relationship between the average SFC-INT and average internal functional connectivity (FC-INT) across modules. The left panel represents Structural Modules, and the right panel represents functional modules. The x-axis shows the average SFC-INT across Modules, and the y-axis shows the average FC-INT across modules. Each point represents a cognitive task, with colors matching the same tasks in both panels.

**Figure S3:**
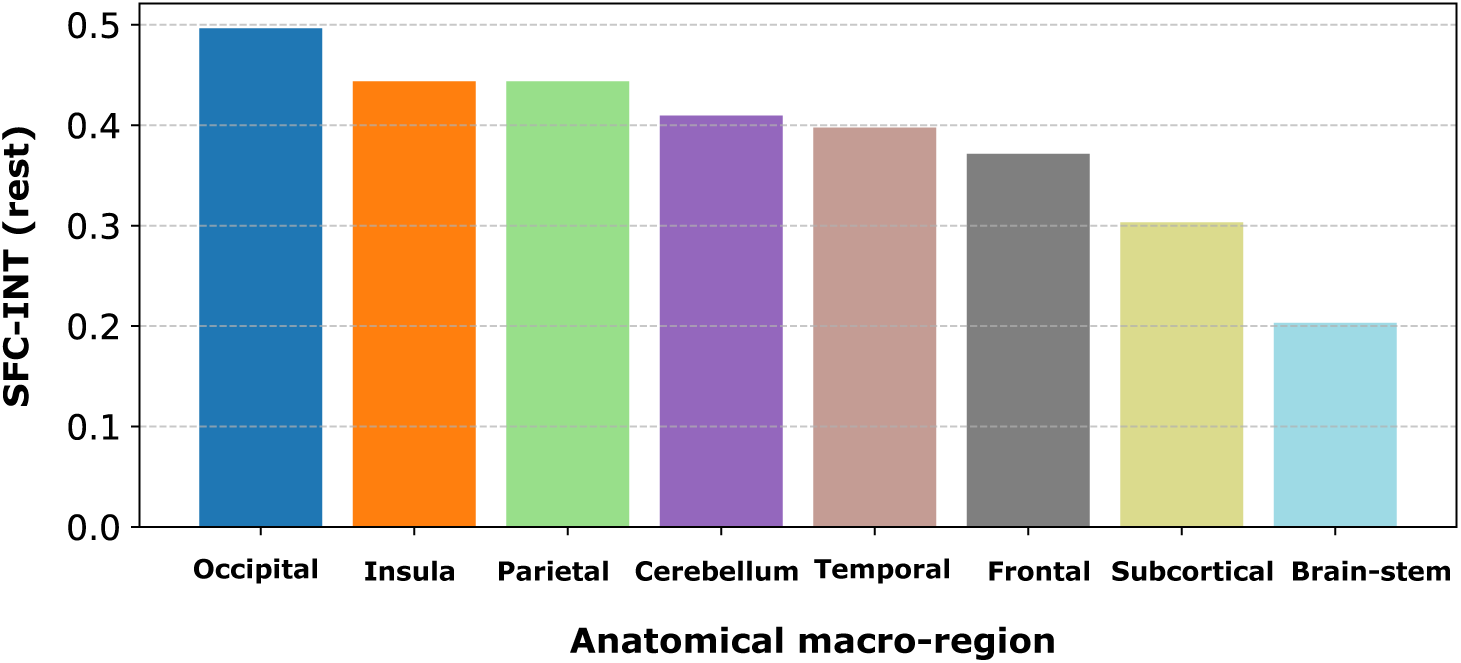
SFC-INT at resting-state across different anatomical macro-regions.

## Footnotes

1 RL: right-left, AP: anterior-posterior, FH: feet-head

