## Supplementary figures and images for "Modular coupling of structure-function reveals network integration (rather than segregation) as the key mechanism for cognitive task discrimination"

### SUPPLEMENTAL FIGURES

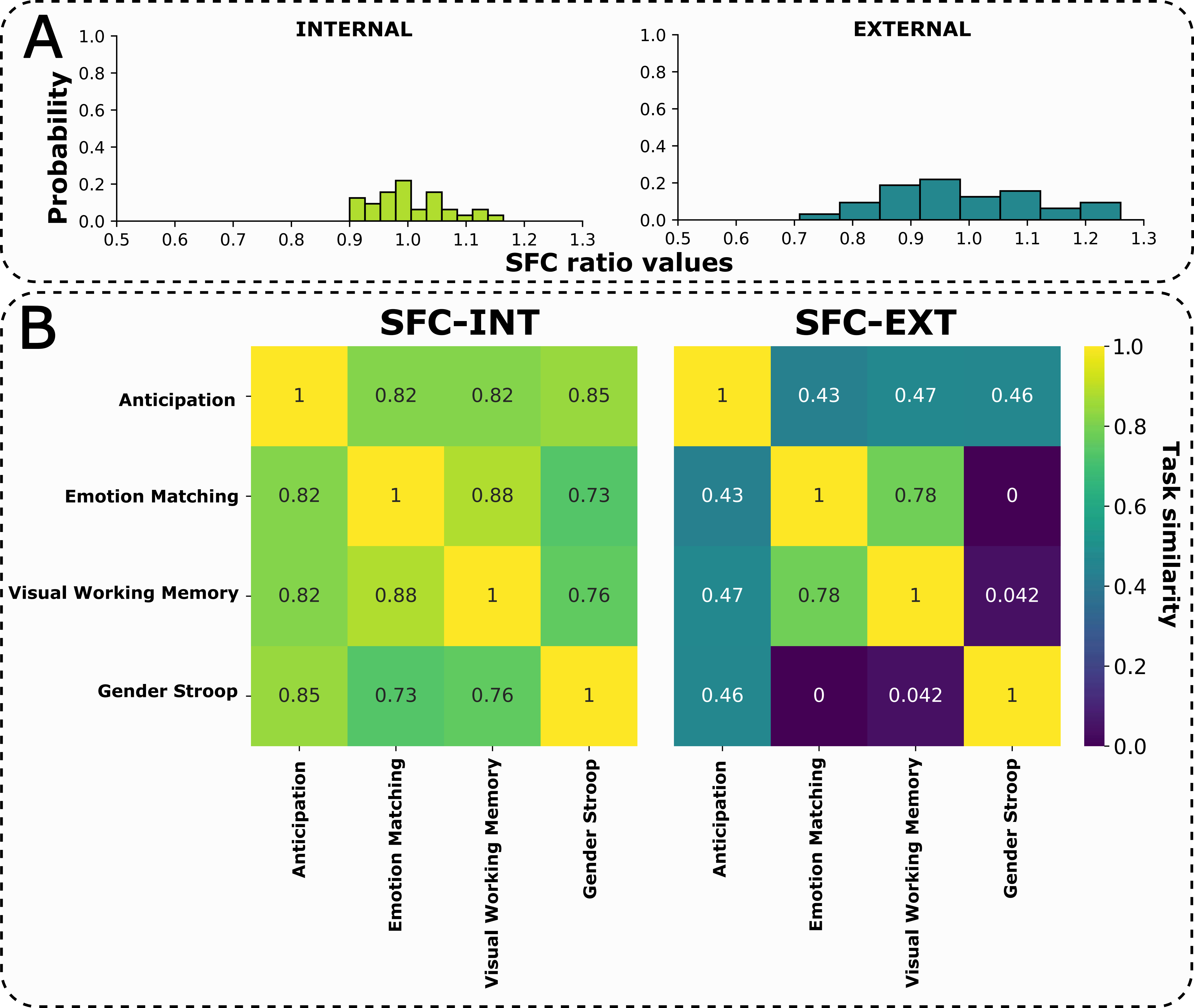

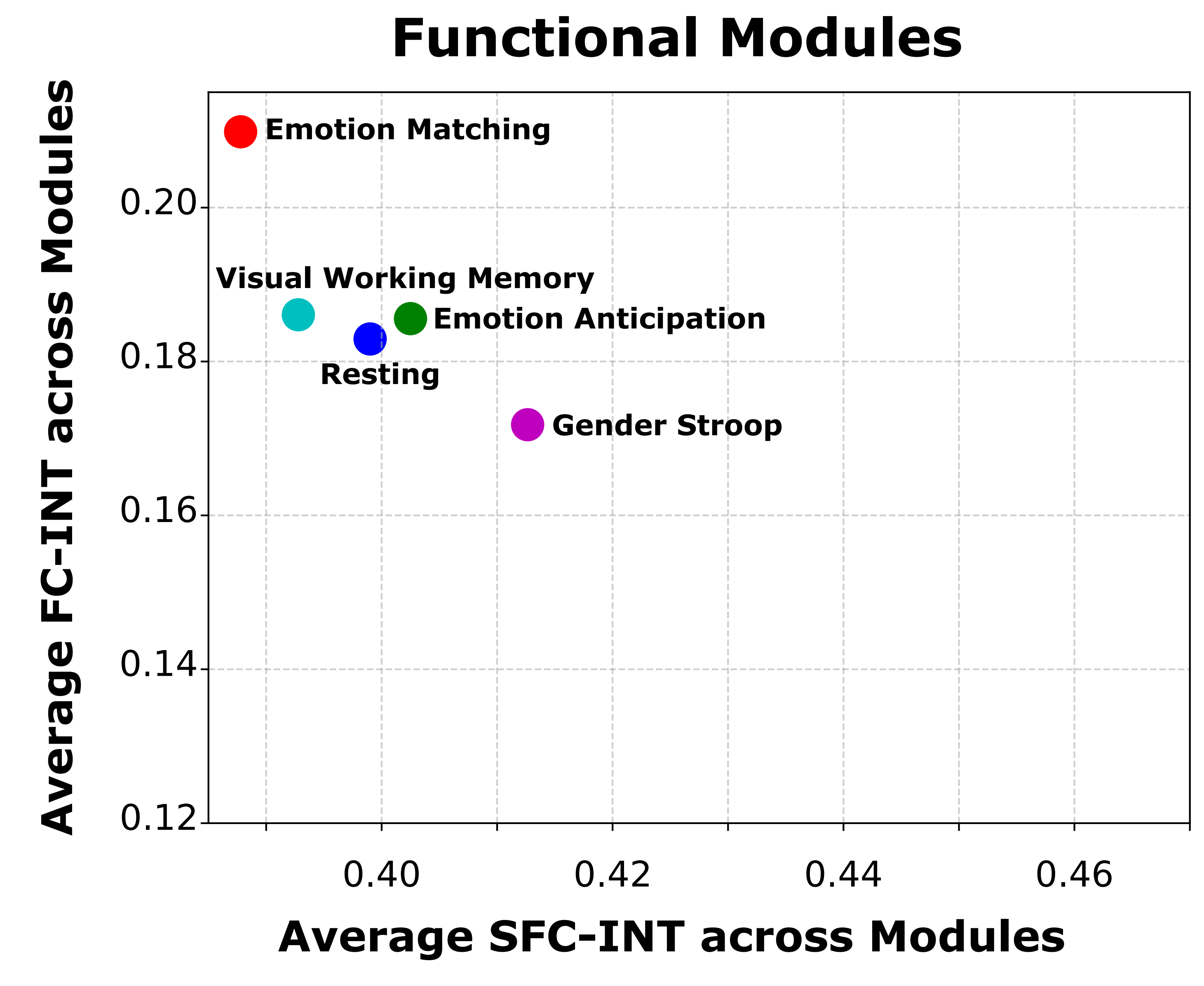

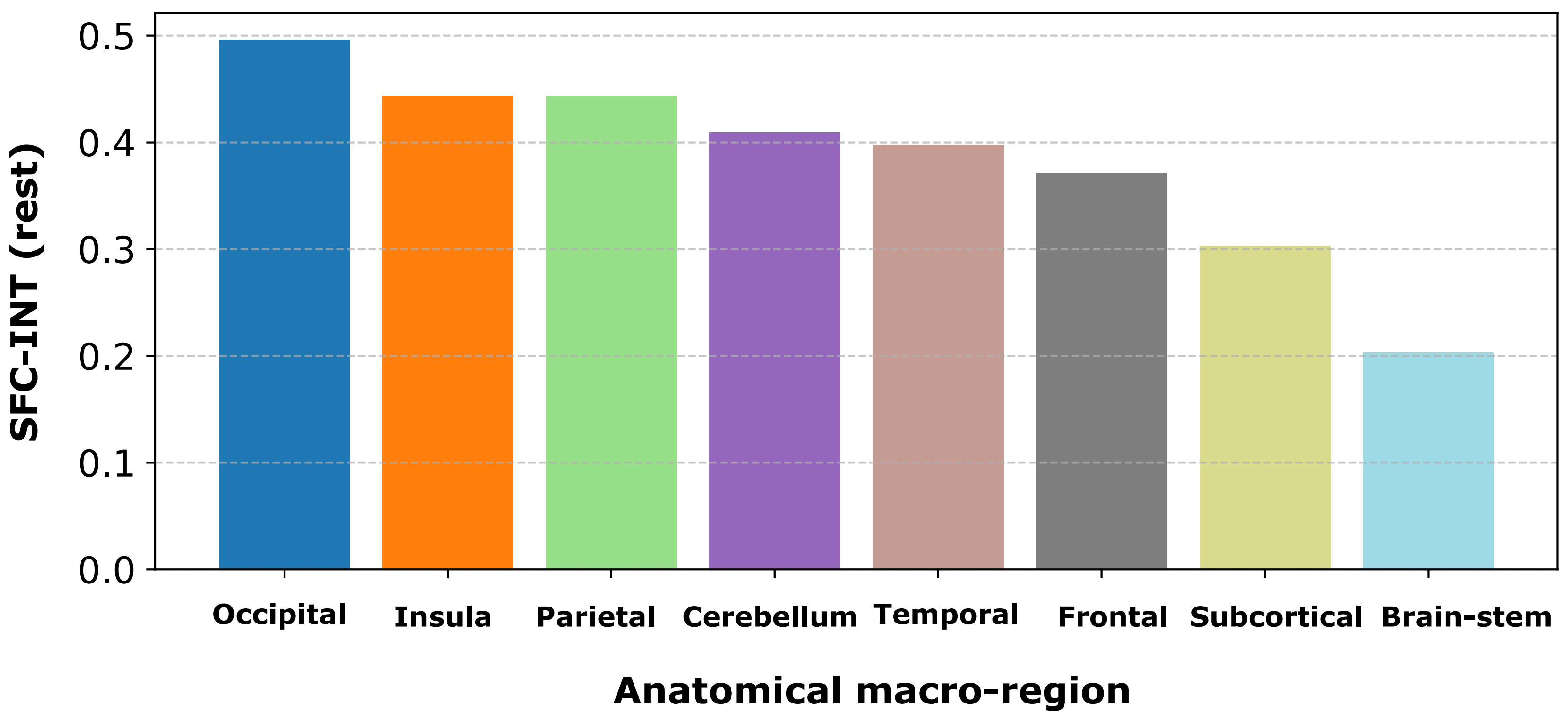
